# DKK1 and CKAP4 expression is associated with cervical lymph node metastasis in tongue squamous cell carcinoma

**DOI:** 10.64898/2026.05.29.26354440

**Authors:** Hironori Fujita, Osamu Takahashi, Naomi Yada, Jumpei Tanaka, Kazuya Haraguchi, Masahiko Morioka, Tatsuki Yaginuma, Masaaki Sasaguri, Shoichiro Kokabu, Manabu Habu

## Abstract

**Objective:** To identify Dickkopf-1 (DKK1) as a prognostically relevant candidate in head and neck squamous cell carcinoma and to evaluate whether DKK1 and cytoskeleton-associated protein 4 (CKAP4) expression is associated with cervical lymph node metastasis in tongue squamous cell carcinoma (TSCC).

**Methods:** DKK1 was screened using the Human Protein Atlas Pathology Atlas. Immunohistochemical expression of DKK1 and CKAP4 was examined in 54 patients with primary TSCC (cT1-4N0) treated surgically between 2015 and 2020. Nine cases were excluded because of insufficient tissue blocks or inadequate staining quality, leaving 45 evaluable cases. Associations with delayed cervical lymph node metastasis were assessed together with conventional clinicopathological factors, including infiltrative growth pattern (INF) and pathological depth of invasion (pDOI).

**Results:** In public database analysis, high DKK1 expression was associated with poorer overall survival in head and neck squamous cell carcinoma. In the TSCC cohort, pDOI ≥5 mm and INF pattern c were significantly associated with cervical lymph node metastasis. Positive DKK1 and CKAP4 expression were also significantly associated with cervical lymph node metastasis. Furthermore, combined DKK1/CKAP4 positivity, when incorporated with INF and pDOI, provided additional risk stratification, and cases with all 3 factors showed a markedly increased likelihood of cervical lymph node metastasis.

**Conclusions:** Expression of DKK1 and CKAP4 was associated with cervical lymph node metastasis in TSCC. Combined assessment of DKK1/CKAP4 expression with INF and pDOI may improve pathological risk stratification and may help identify patients who require closer neck evaluation and postoperative management.

## Introduction

Most malignant tumors arising in the oral cavity are oral squamous cell carcinomas (OSCCs), and the tongue is one of the most common and clinically important subsites of oral cavity squamous cell carcinoma [1]. In tongue squamous cell carcinoma (TSCC), the presence of cervical lymph node metastasis is a major determinant of survival and an important factor in treatment decision-making [2]. However, accurate prediction of cervical lymph node metastasis based on the initial presentation or histopathological findings of the primary tumor remains challenging.

Several clinicopathological parameters have been reported to be useful for estimating the risk of cervical lymph node metastasis in OSCC. Among them, pathological depth of invasion (pDOI) has consistently been identified as a robust predictor of nodal metastasis, and histopathological features at the invasive front, including infiltrative growth pattern (INF) or related patterns of invasion, have also been associated with nodal relapse and poor outcome [3-6]. However, these conventional parameters alone do not fully stratify metastatic risk. Even when neck management is guided by DOI-based criteria, a substantial proportion of patients do not harbor occult nodal disease, underscoring the limitations of current pathological indicators and the need for novel biomarkers that enable more precise risk assessment [6].

Dickkopf-1 (DKK1) is a secreted protein best known as an antagonist of the canonical Wnt/β-catenin signaling pathway, acting mainly through low-density lipoprotein receptor-related proteins 5 and 6 (LRP5/6) [7]. Wnt signaling has been widely implicated in tumor cell proliferation, invasion, metastasis, and prognosis, and is thought to play an important role in a variety of malignancies, including head and neck squamous cell carcinoma [8, 9]. On the basis of this biological background, DKK1 was initially considered a potential tumor-suppressive molecule. However, DKK1 was later found to be overexpressed in lung and esophageal carcinomas and to be associated with poor prognosis [10]. In addition, anti-DKK1 antibody was shown to inhibit cancer cell growth and invasion, suggesting that DKK1 may contribute to tumor progression rather than functioning solely as a canonical Wnt inhibitor [11]. Furthermore, Kimura et al. identified cytoskeleton-associated protein 4 (CKAP4) as a receptor for DKK1 and demonstrated that DKK1 activates the PI3K–AKT pathway through CKAP4 to promote cancer cell proliferation [12]. These findings suggest that, apart from its inhibitory effect on canonical Wnt/β-catenin signaling, DKK1 may also participate in tumor development and malignant progression through an alternative signaling route mediated by CKAP4.

In this study, we first identified DKK1 as a prognostically relevant candidate through analysis of a public head and neck squamous cell carcinoma dataset. We then evaluated the expression of DKK1 and CKAP4 immunohistochemically in tongue squamous cell carcinoma specimens and examined their associations with cervical lymph node metastasis. Finally, we assessed whether combining DKK1/CKAP4 co-expression with conventional pathological parameters, including infiltrative growth pattern (INF) and pathological depth of invasion (pDOI), could improve risk stratification for cervical lymph node metastasis.

## Materials and methods

### Public database analysis

Candidate proteins associated with prognosis in head and neck squamous cell carcinoma were screened using the Human Protein Atlas pathology atlas [13]. Data on mRNA expression, overall survival, and follow-up in 499 patients with head and neck squamous cell carcinoma from The Cancer Genome Atlas were then collected and analyzed [14]. Based on these data, DKK1 was selected as a candidate molecule for further investigation.

### Patients

This retrospective study included 54 patients with primary tongue squamous cell carcinoma (clinical stage cT1-4N0) who underwent surgery under general anesthesia at our institution between 2015 and 2020 and were followed for at least 30 months after surgery. The patients ranged in age from 22 to 89 years (mean ± standard deviation, 63.5 ± 15.7 years) and included 33 men and 21 women. We retrospectively investigated the associations of DKK1 and CKAP4 expression, assessed by immunohistochemistry, with delayed cervical lymph node metastasis, together with clinical and histopathological findings. Written informed consent was obtained from all patients before surgery. This study was approved by the Research Ethics Committee of Kyushu Dental University (approval no. 20-32).

### Clinicopathological evaluation

Clinical T classification was T1 in 20 patients, T2 in 24, T3 in 8, and T4 in 2, according to the Union for International Cancer Control TNM classification, 8th edition. Histopathological evaluation of the resected specimens included histological differentiation, infiltrative growth pattern (INF), lymphatic vessel invasion (Ly), venous invasion (V), perineural invasion (Pn), pathological depth of invasion (pDOI), and the presence or absence of delayed cervical lymph node metastasis.

### Immunohistochemistry

Expression of DKK1 and CKAP4 was evaluated immunohistochemically in resected specimens. The specimens were fixed in 10% neutral buffered formalin, embedded in paraffin, and sectioned at a thickness of 4 μm. After deparaffinization and rehydration, endogenous peroxidase activity was blocked using KPL Blocking Solution Concentrate (SeraCare, Milford, MA, USA). Serum blocking was then performed using the VECTASTAIN Elite ABC Kit (Vector Laboratories, Burlingame, CA, USA). The sections were incubated overnight at 4°C with anti-DKK1 antibody (1:80 dilution; Santa Cruz Biotechnology, Dallas, TX, USA) or anti-CKAP4 antibody (1:80 dilution; Enzo Life Sciences, Farmingdale, NY, USA). After incubation with the primary antibodies, the sections were incubated for 30 min at room temperature with the biotinylated universal anti-mouse/rabbit IgG secondary antibody included in the VECTASTAIN Elite ABC Kit.Immunoreactivity was then visualized using the KPL DAB Reagent Set (SeraCare), followed by counterstaining with hematoxylin. Immunohistochemical positivity for DKK1 or CKAP4 was defined as staining in more than 20% of tumor cells. Staining results were independently evaluated by three investigators who were blinded to the clinical outcomes. Of the 54 cases, 9 were excluded from the immunohistochemical analysis because of insufficient tissue blocks or inadequate staining quality. Therefore, 45 cases were included in the evaluable immunohistochemical cohort.

### Statistical analysis

All statistical analyses were performed using EZR [15], a graphical user interface for R designed for medical statistics. In the public database analysis, overall survival was evaluated using the Kaplan-Meier method, and differences between groups were assessed using the log-rank test. In the clinicopathological analyses of the tongue squamous cell carcinoma cohort, associations between clinicopathological variables or immunohistochemical findings and delayed cervical lymph node metastasis were evaluated by univariate logistic regression analysis. Odds ratios (ORs) and 95% confidence intervals (CIs) were calculated. A p value of < 0.05 was considered statistically significant.

## Results

### DKK1 as a prognostically relevant candidate in head and neck squamous cell carcinoma

To identify candidate molecules associated with poor prognosis in head and neck squamous cell carcinoma, we first performed an in silico screening using the Human Protein Atlas pathology atlas based on the TCGA cohort (Figure 1A). Among the top unfavorable prognostic genes listed in this dataset, DKK1 was selected as a candidate for further investigation (Figure 1B). Kaplan–Meier survival analysis of the TCGA-based head and neck squamous cell carcinoma cohort showed that high DKK1 expression was associated with poorer overall survival than low DKK1 expression (Figure 1C). In addition, the survival scatter plot demonstrated the distribution of individual cases according to DKK1 expression level and follow-up status, further supporting the association between elevated DKK1 expression and unfavorable clinical outcome (Figure 1D).

**Figure 1.**
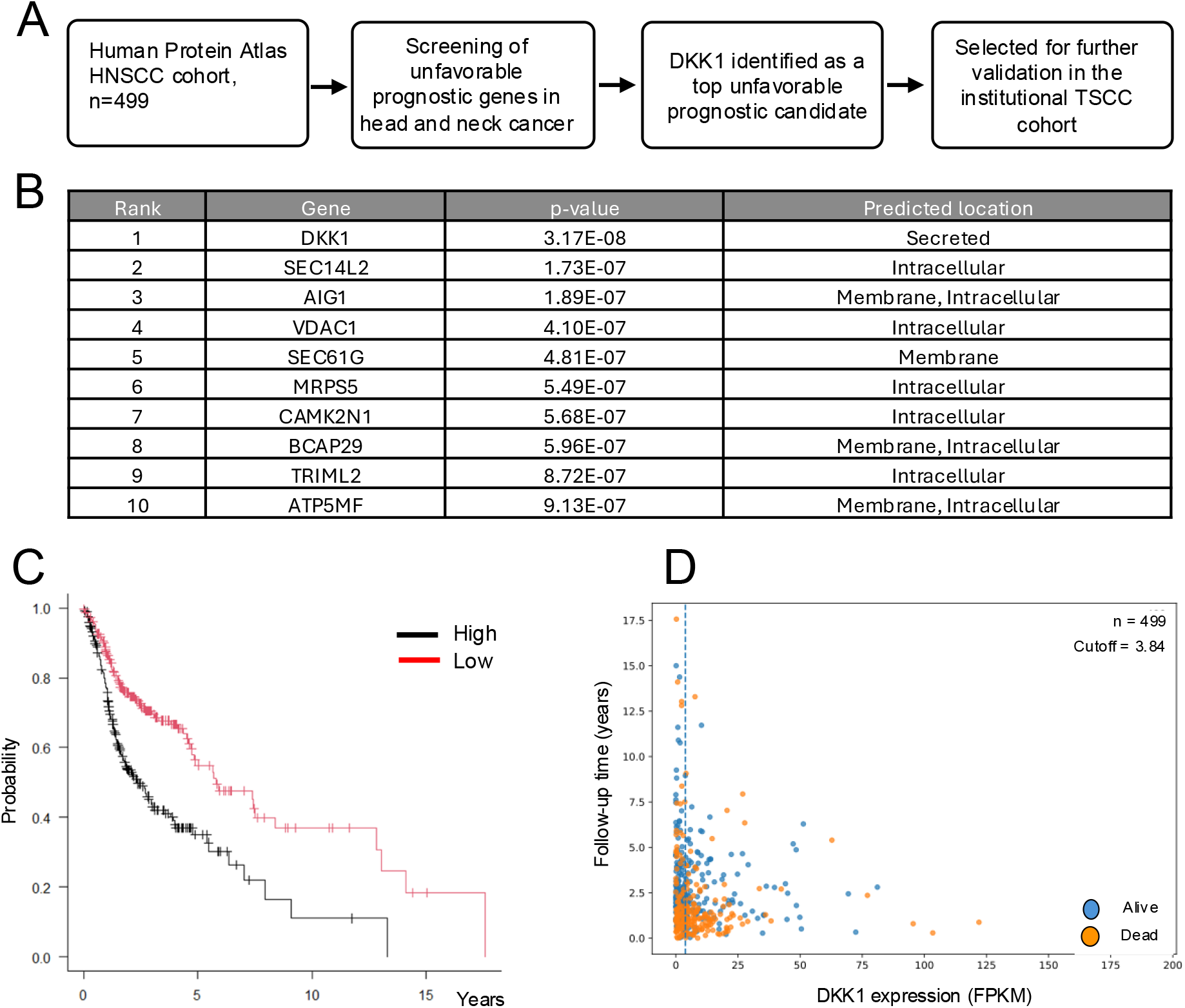
In silico identification of DKK1 as a prognostic candidate. (A) Workflow of candidate selection using the Human Protein Atlas database. (B) Top 10 unfavorable prognostic genes in the head and neck squamous cell carcinoma dataset. (C) Kaplan–Meier survival analysis according to DKK1 expression in the TCGA-based cohort. (D) Survival scatter plot showing DKK1 expression and follow-up status in individual cases.

### DKK1 and CKAP4 expression were associated with cervical lymph node metastasis in tongue squamous cell carcinoma

Because the primary focus of this study was tongue squamous cell carcinoma, all subsequent analyses were performed using a tongue squamous cell carcinoma cohort. The study cohort consisted of 54 patients, including 33 without cervical lymph node metastasis and 21 with cervical lymph node metastasis (Table 1). Clinical T classification was distributed as follows: T1 in 20 patients, T2 in 24, T3 in 8, and T4 in 2 (Table 1). We next examined the associations between conventional clinicopathological factors and cervical lymph node metastasis in this cohort. A pDOI of ≥5 mm was significantly associated with cervical lymph node metastasis compared with a pDOI of ≤5 mm (OR, 10.64; 95% CI, 2.53–44.80; p = 0.001). Likewise, an INF pattern of c was significantly associated with cervical lymph node metastasis compared with INF pattern a/b (OR, 8.40; 95% CI, 2.19–32.15; p = 0.001) (Table 2).

**Table 1.** Clinicopathological characteristics of the study cohort.

| pDOI and cervical lymph node metastasis |  |  |  |  |  |  |
| --- | --- | --- | --- | --- | --- | --- |
| Variable | Category | LN (–), n | LN (+), n | OR | 95% CI | p-value |
| pDOI | ≤5 mm | 27 | 3 | Ref. |  |  |
|  | ≥5 mm | 11 | 13 | 10.64 | 2.53–44.80 | 0.001 |
| INF pattern and cervical lymph node metastasis |  |  |  |  |  |  |
| Variable | Category | LN (–), n | LN (+), n | OR | 95% CI | p-value |
| INF pattern | a / b | 28 | 4 | Ref. |  |  |
|  | c | 10 | 12 | 8.40 | 2.19–32.15 | 0.001 |
| *Odds ratios were calculated against the first-listed reference category.* |  |  |  |  |  |  |

**Table 2.**
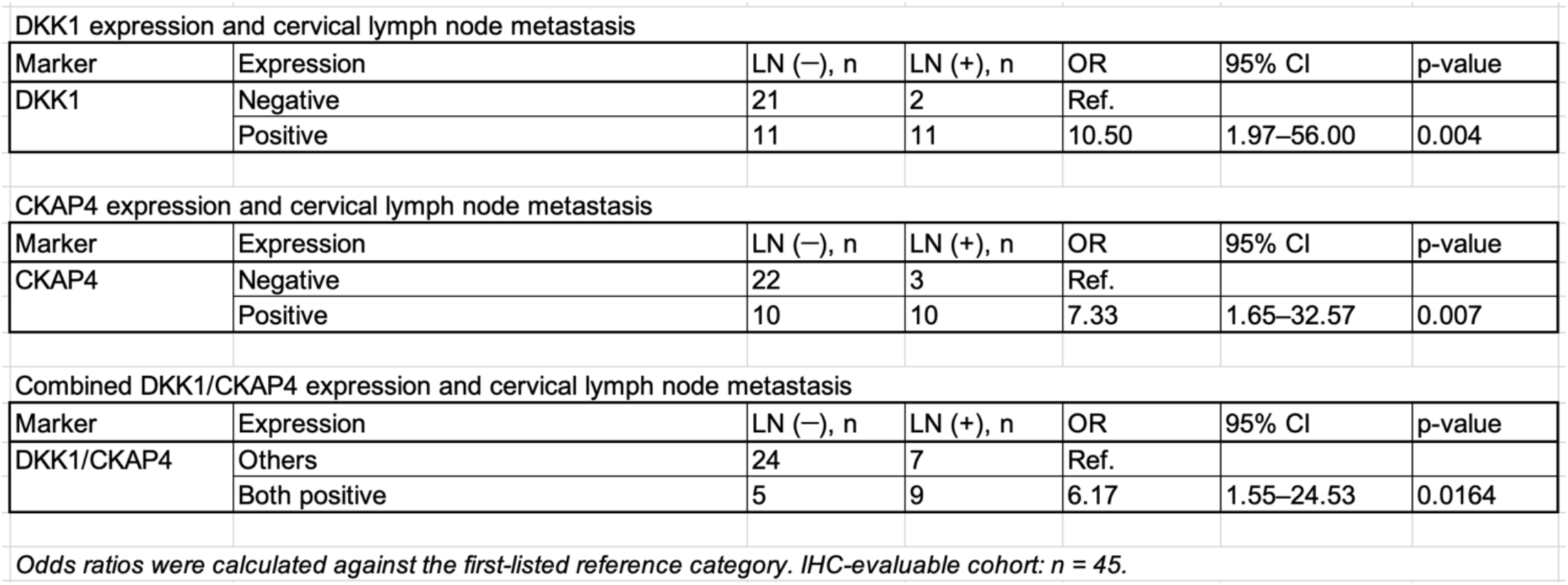
Associations between clinicopathological factors and cervical lymph node metastasis.

| DKK1 expression and cervical lymph node metastasis |  |  |  |  |  |  |
| --- | --- | --- | --- | --- | --- | --- |
| Marker | Expression | LN (–), n | LN (+), n | OR | 95% CI | p-value |
| DKK1 | Negative | 21 | 2 | Ref. |  |  |
|  | Positive | 11 | 11 | 10.50 | 1.97–56.00 | 0.004 |
| CKAP4 expression and cervical lymph node metastasis |  |  |  |  |  |  |
| Marker | Expression | LN (–), n | LN (+), n | OR | 95% CI | p-value |
| CKAP4 | Negative | 22 | 3 | Ref. |  |  |
|  | Positive | 10 | 10 | 7.33 | 1.65–32.57 | 0.007 |
| Combined DKK1/CKAP4 expression and cervical lymph node metastasis |  |  |  |  |  |  |
| Marker | Expression | LN (–), n | LN (+), n | OR | 95% CI | p-value |
| DKK1/CKAP4 | Others | 24 | 7 | Ref. |  |  |
|  | Both positive | 5 | 9 | 6.17 | 1.55–24.53 | 0.0164 |

We then evaluated the protein expression of DKK1 and its receptor candidate CKAP4 by immunohistochemistry in tongue squamous cell carcinoma specimens. Representative immunohistochemical staining of DKK1 and CKAP4 is shown in Figure 2. Of the 54 cases, 45 were evaluable for immunohistochemical analysis, whereas 9 cases were excluded because of insufficient tissue blocks or inadequate staining quality. In this evaluable cohort, positive DKK1 expression was significantly associated with cervical lymph node metastasis (OR, 10.50; 95% CI, 1.97–56.00; p = 0.004). Positive CKAP4 expression was also significantly associated with cervical lymph node metastasis (OR, 7.33; 95% CI, 1.65–32.57; p = 0.007). Furthermore, combined positivity for DKK1 and CKAP4 was significantly associated with cervical lymph node metastasis compared with the remaining cases (OR, 6.17; 95% CI, 1.55–24.53; p = 0.0164) (Table 3).

**Table 3.** Associations between DKK1 and CKAP4 expression and cervical lymph node metastasis.

| DKK1 expression and cervical lymph node metastasis |  |  |  |  |  |  |
| --- | --- | --- | --- | --- | --- | --- |
| Marker | Expression | LN (–), n | LN (+), n | OR | 95% CI | p-value |
| DKK1 | Negative | 21 | 2 | Ref. |  |  |
|  | Positive | 11 | 11 | 10.50 | 1.97–56.00 | 0.004 |
| CKAP4 expression and cervical lymph node metastasis |  |  |  |  |  |  |
| Marker | Expression | LN (–), n | LN (+), n | OR | 95% CI | p-value |
| CKAP4 | Negative | 22 | 3 | Ref. |  |  |
|  | Positive | 10 | 10 | 7.33 | 1.65–32.57 | 0.007 |
| Combined DKK1/CKAP4 expression and cervical lymph node metastasis |  |  |  |  |  |  |
| Marker | Expression | LN (–), n | LN (+), n | OR | 95% CI | p-value |
| DKK1/CKAP4 | Others | 24 | 7 | Ref. |  |  |
|  | Both positive | 5 | 9 | 6.17 | 1.55–24.53 | 0.0164 |
| Odds ratios were calculated against the first-listed reference category. IHC-evaluable cohort: n = 45. |  |  |  |  |  |  |

**Figure 2.**
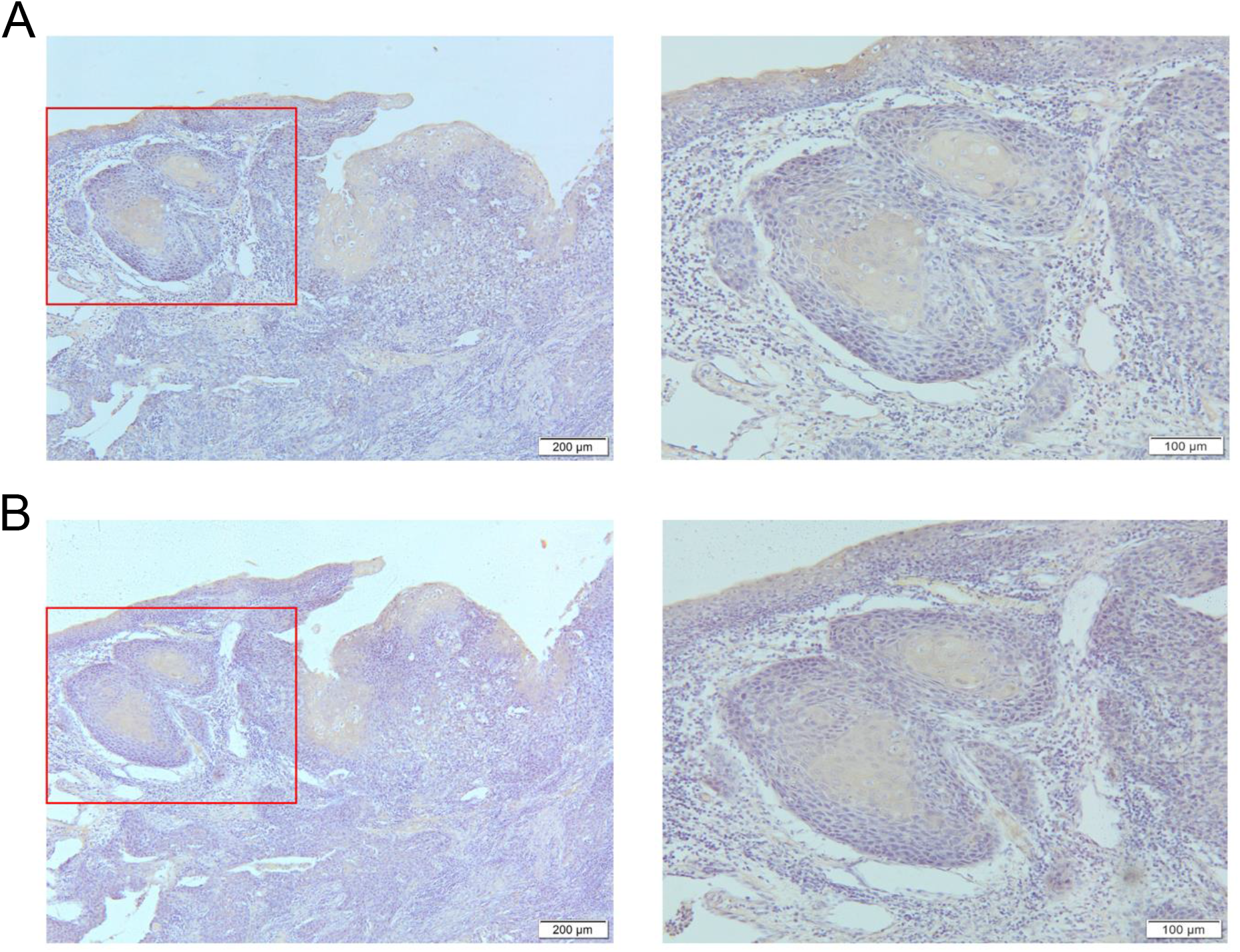
Representative immunohistochemical staining of DKK1 and CKAP4. (A) Representative DKK1 staining at low magnification (left) and high magnification (right). The right panel shows a magnified view of the boxed area in the left panel. Scale bars, 200 μm (left) and 100 μm (right). (B) Representative CKAP4 staining at low magnification (left) and high magnification (right). The right panel shows a magnified view of the boxed area in the left panel. Scale bars, 200 μm (left) and 100 μm (right).

### Combined assessment of DKK1/CKAP4 co-expression, INF pattern, and pDOI provided additional stratification for cervical lymph node metastasis

Finally, we constructed a risk stratification model for cervical lymph node metastasis based on INF, pDOI, and DKK1/CKAP4 co-expression. Cases with 0–1 risk factors were classified as low risk, those with 2 risk factors as intermediate risk, and those with all 3 risk factors—INF pattern c, pDOI ≥5 mm, and DKK1/CKAP4 co-expression—as high risk. Compared with the low-risk group, the intermediate-risk group showed increased odds of cervical lymph node metastasis (OR, 9.78; 95% CI, 0.96–99.94; p = 0.0470), whereas the high-risk group showed a markedly elevated risk (OR, 176.00; 95% CI, 9.81–3158.95; p < 0.0001) (Table 4). These findings suggest that combined assessment of DKK1/CKAP4 co-expression with INF pattern and pDOI may provide a useful framework for stratifying the risk of cervical lymph node metastasis in tongue squamous cell carcinoma.

**Table 4.** Risk stratification based on INF, pDOI, and DKK1/CKAP4 co-expression.

| Risk group | Definition | LN (–), n | LN (+), n | OR vs low | 95% CI | p-value |
| --- | --- | --- | --- | --- | --- | --- |
| Low risk | 0–1 risk factors | 22 | 1 | Ref. |  |  |
| Intermediate risk | 2 risk factors | 9 | 4 | 9.78 | 0.96–99.94 | 0.047 |
| High risk | INF c + pDOI ≥5 mm + DKK1/CKAP4 (+) | 1 | 8 | 176 | 9.81–3158.95 | <0.0001 |

*Odds ratios were calculated using the low-risk group as the reference.*

## Discussion

In the present study, we first identified DKK1 as a prognostically relevant candidate through in silico analysis of a public head and neck squamous cell carcinoma dataset and subsequently examined its clinicopathological significance in tongue squamous cell carcinoma. Immunohistochemical analysis demonstrated that both DKK1 and CKAP4 expression were significantly associated with cervical lymph node metastasis in tongue squamous cell carcinoma. Moreover, combined positivity for DKK1 and CKAP4 was also associated with nodal metastasis, and incorporation of DKK1/CKAP4 co-expression into a model including INF and pDOI provided additional stratification for cervical lymph node metastasis. Collectively, these findings suggest that the DKK1/CKAP4 axis may have clinicopathological relevance in identifying tongue squamous cell carcinoma with higher metastatic potential.

The association of DKK1 and CKAP4 with cervical lymph node metastasis observed in the present study is compatible with previous evidence indicating that DKK1 may have protumorigenic effects beyond its canonical role as a Wnt antagonist and that CKAP4 functions as a receptor mediating DKK1-driven PI3K– AKT signaling [10-12]. These findings raise the possibility that the DKK1/CKAP4 axis is involved in the acquisition of a more aggressive phenotype in tongue squamous cell carcinoma.

From a clinical perspective, our findings suggest that DKK1 and CKAP4 may serve as adjunctive biomarkers in the pathological assessment of metastatic risk in tongue squamous cell carcinoma. In particular, the combined model incorporating DKK1/CKAP4 co-expression with INF and pDOI identified a subgroup with a markedly increased likelihood of cervical lymph node metastasis. Although these markers are not intended to replace established pathological indicators, they may provide an additional layer of information for identifying patients who warrant closer evaluation of the neck and more careful postoperative management.

Several limitations of this study should be acknowledged. First, this was a retrospective study conducted at a single institution with a relatively small sample size. Second, immunohistochemical evaluation was not possible in 9 cases because of insufficient tissue blocks or inadequate staining quality, which may have introduced selection bias. Third, although the initial in silico screening was performed using a public head and neck squamous cell carcinoma dataset, the subsequent clinicopathological validation was restricted to tongue squamous cell carcinoma. While this disease-focused validation represents a strength of the present study, caution is warranted in extrapolating the findings to other head and neck subsites. Finally, this study was observational in nature, and no functional experiments were performed to directly determine whether the DKK1/CKAP4 axis contributes to invasion or metastasis in tongue squamous cell carcinoma. Therefore, further validation in larger, independent cohorts and mechanistic studies will be necessary.

In conclusion, expression of DKK1 and CKAP4 was associated with cervical lymph node metastasis in tongue squamous cell carcinoma, and their combined assessment with INF and pDOI provided additional stratification. These findings suggest that the DKK1/CKAP4 axis may complement conventional pathological evaluation.

## Data Availability

The data that support the findings of this study are available from the corresponding author upon reasonable request. Publicly available data used for the in silico analysis are available through the Human Protein Atlas Pathology Atlas.

https://www.proteinatlas.org/humanproteome/cancer/head%2Band%2Bneck%2Bsquamous%2Bcell%2Bcarcinoma

https://v21.proteinatlas.org/ENSG00000107984-DKK1/pathology

---

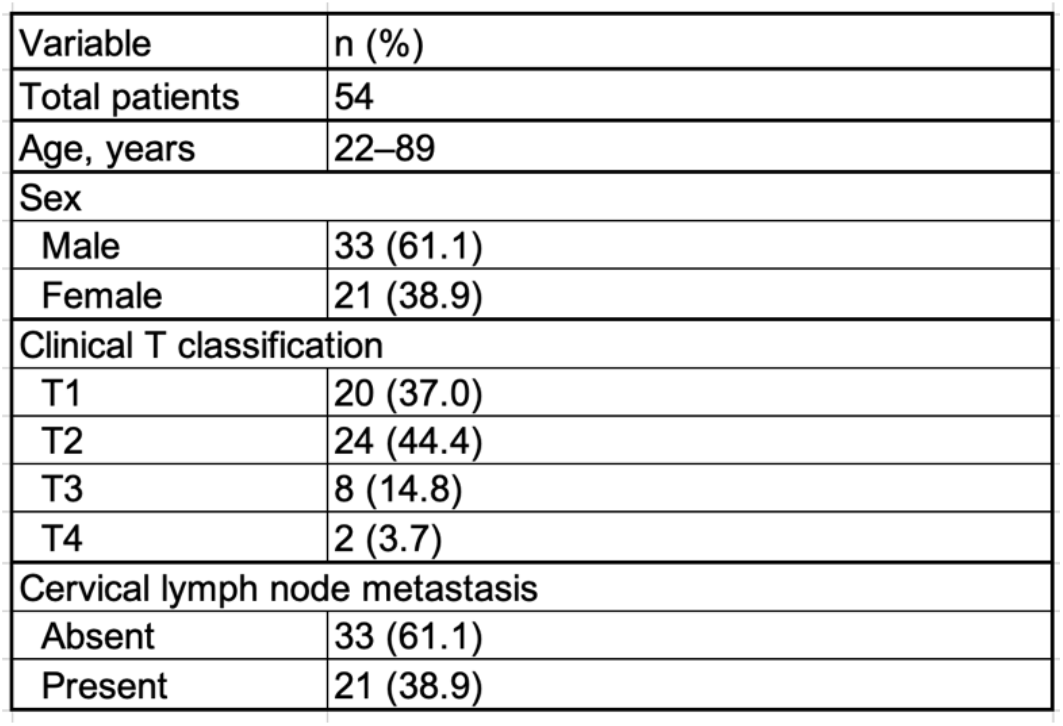

## Author Contributions

H.F., M.H., and S.K. conceived and designed the study. H.F., O.T., J.T., K.T., K.H., M.M., and T.Y. collected the clinical data and performed the immunohistochemical analyses. H.F. and S.K. analyzed the data. H.F. and N.Y. provided pathological evaluation and interpretation. H.F., O.T., and S.K. drafted the manuscript. All authors reviewed the manuscript and approved the final version.

## Ethics Statement

This study was approved by the Research Ethics Committee of Kyushu Dental University (approval no. 20-32). Written informed consent was obtained from all patients before surgery.

## Data Availability Statement

The data that support the findings of this study are available from the corresponding author upon reasonable request.

